## Supplemental Information for "Allelic strengths of encephalopathy-associated *UBA5* variants correlate between *in vivo* and *in vitro* assays"

### Case report

The reported proband is a boy with axial hypotonia, generalized dystonia, lower extremity spasticity, global developmental delay, esotropia and failure to thrive. An electroencephalogram (EEG) showed multifocal epileptiform discharges in drowsiness and sleep. However, he has not had seizures. Two magnetic resonance imaging (MRI) studies at different ages were read as normal, although on review show a slightly thin corpus callosum, posterior periventricular white matter T2 hyperintensity resulting in increased conspicuity of the subcortical U-fibers and widening of the Sylvian fissures (Figure S1). He is not microcephalic. For more information about the clinical record of the proband please contact the corresponding author.

On trio exome sequencing, the proband was found to have compound heterozygous variants in *UBA5*, NM_024818.6:c.169A>G (p.Met57Val) and c.935A>T (p.Gln312Leu). The two variants are in *trans* phase. The p.Met57Val variant has been reported in one individual with DEE44 (Colin *et al*, 2016). The p.Gln312Leu variant has not been previously reported. Neither variant is reported in the gnomAD database v.2.1.1 (https://gnomad.broadinstitute.org). Both variants are predicted to be damaging or probably damaging by multiple pathogenicity prediction tools (Table S3).

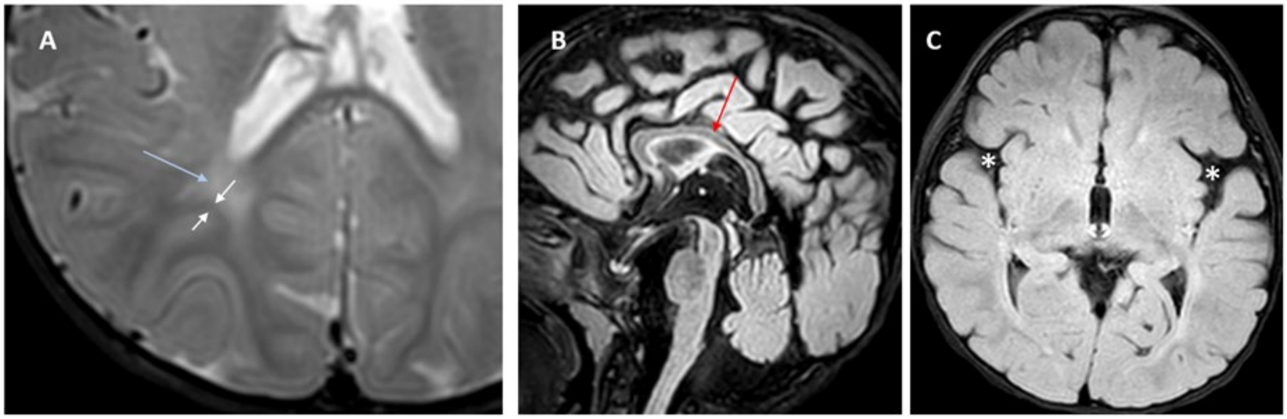


**Figure S1. Brain magnetic resonance imaging (MRI) images**

(A) Axial T2 image showing periventricular T2 hyperintensity (blue arrow) resulting in prominence of the subcortical U-fibers (white arrows).

(B) Sagittal flair showing mild thinning of the corpus callosum (red arrow).

(C) Axial flair image demonstrating widening of the sylvian fissures (white asterisks).

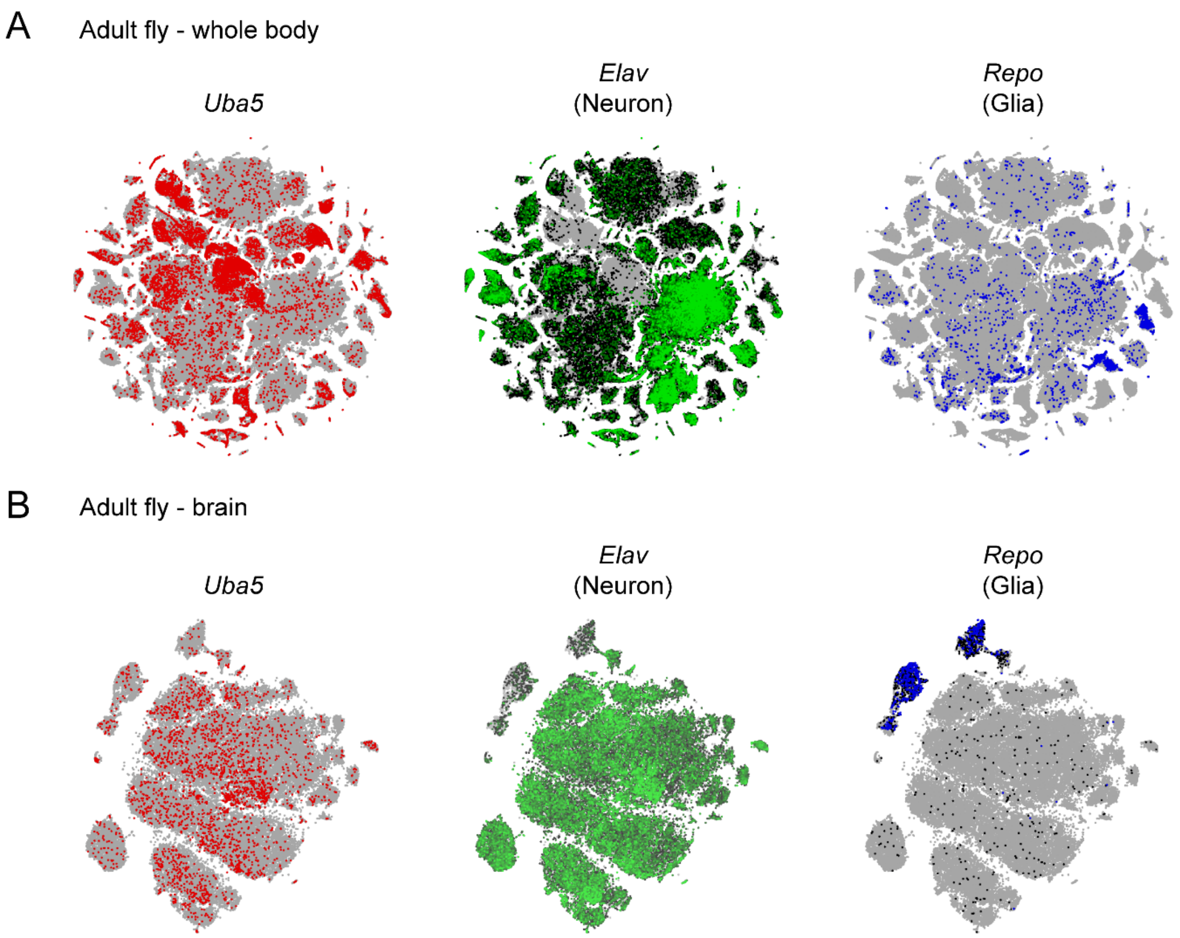


C

| | *Uba5*$^{KO}$/ *Y; UAS-UBA5*$^{Ref}$ | *Uba5*$^{KO}$/ *+; UAS-UBA5*$^{Ref}$ |
|---|---|---|
| *No driver* | Embyonic / L1 lethal | Viable, NOP |
| *Da-Gal4* | Viable, NOP | Viable, NOP |
| *Act-Gal4* | Viable, NOP | Viable, NOP |
| *Lpp-Gal4* | Embyonic / L1 lethal | Viable, NOP |
| *Mef2-Gal4* | Embyonic / L1 lethal | Viable, NOP |
| *Elav-Gal4* | Embyonic / L1 lethal | Viable, NOP |
| *Repo-Gal4* | Embyonic / L1 lethal | Viable, NOP |
| *Elav-Gal4, Repo-Gal4* | Embyonic / L1 lethal | Viable, NOP |

**Figure S2. Single-cell gene expression pattern of *Uba5* and the rescue of *Uba5* mutants by tissue-specific *UBA5* expression**

(A and B) The expression pattern of *Uba5* in whole adult fly (Li *et al*, 2022) (A) and adult brain tissue (Davie *et al*, 2018) (B) revealed by single-cell RNA sequencing profiles. The expression patterns of neuronal marker *Elav* and glial marker *Repo* are also shown.

(C) The lethality of *Uba5* hemizygous mutants is rescued by ubiquitous expression, but not by any tissue-specific expression of human *UBA5* cDNA. Overexpression of *UBA5* in *Uba5* heterozygous mutants does not cause any obvious phenotype.

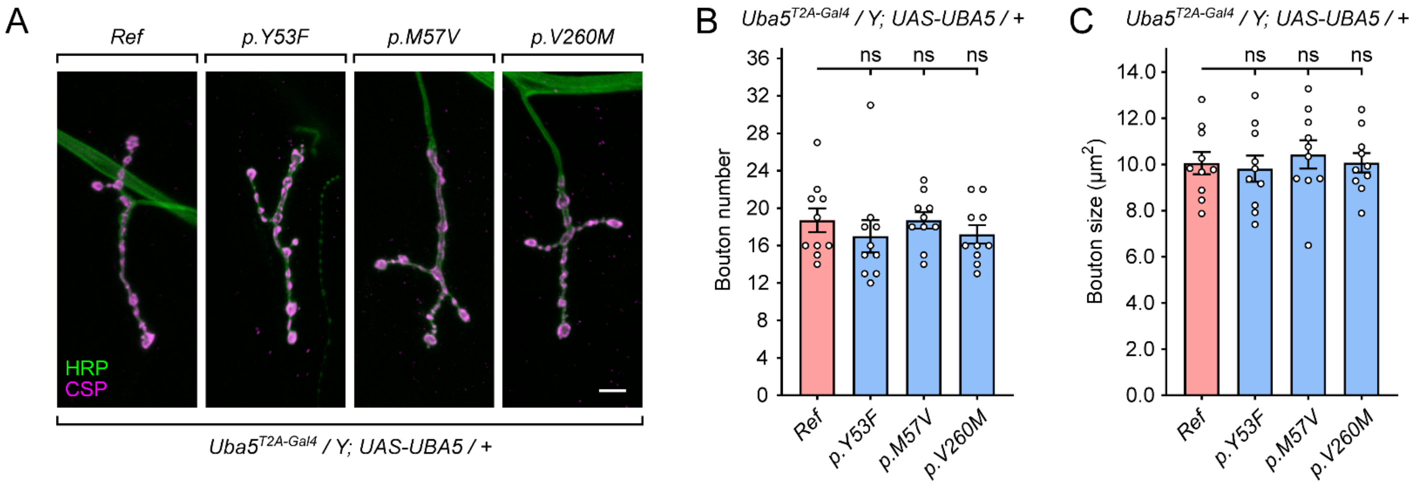


**Figure S3. The Group II *UBA5* variants do not cause obvious synaptic growth defects**

(A) Images of NMJ4 in segments A2-A4 stained with anti-horseradish peroxidase (HRP) and anti-cysteine string protein (CSP) in humanized flies expressing reference *UBA5* or Group II variants. Scale bar, 10 μm.

(B and C) Quantification of the bouton number (B) and bouton size (C) of NMJs. Results are presented as means ± SEM. Statistical analyses were performed via two-sided, unpaired Student's t-test. ns, not significant.

**Table S1. Summary of genotypes of the reported cases**

| References | Family | Allele #1 | Allele #2 |
|---|---|---|---|
| Colin, *et al.*, 2016 (Colin *et al.*, 2016) | A | p.Ala371Thr (IA†) | p.Gln302* |
| | B | p.Ala371Thr (IA) | p.Lys324Asnfs*14 |
| | C | p.Asp389Tyr (IA) | p.Val260Met (II) |
| | D | p.Met57Val (II) | p.Gly168Glu (III) |
| Muona, *et al.*, 2016 (Muona *et al*, 2016) | A | p.Ala371Thr (IA) | p.Arg55His (III) |
| | B | p.Ala371Thr (IA) | p.Tyr285* |
| | C, E | p.Ala371Thr (IA) | p.Arg188* |
| | D | p.Ala371Thr (IA) | p.Arg61* |
| Arnadottir, *et al.*, 2017 (Arnadottir *et al*, 2017) | | p.Ala371Thr (IA) | p.Ala288= (splicing variant) |
| Daida, *et al.*, 2018 (Daida *et al*, 2018) | | p.Tyr72Cys (IB) | Deletion |
| Mignon-Ravix, *et al.*, 2018 (Mignon-Ravix *et al*, 2018) | | p.Tyr53Phe (II)†† | p.Tyr53Phe (II) |
| Low, *et al.*, 2019 (Low *et al*, 2019) | | p.Asp389Gly (IA) | Deletion |
| Briere, *et al.*, 2021 (Briere *et al*, 2021) | A | p.Ala371Thr (IA) | p.Cys303Arg (III) |
| | B | p.Ala371Thr (IA) | p.Arg188* |
| | C | p.Ala371Thr (IA) | p.Leu254Pro (III) |
| | D | p.Ala371Thr (IA) | p.Cys303Arg (III) |
| This study | | p.Met57Val (II) | p.Gln312Leu (IB) |

† The variant classification using fly phenotypic assays (results shown in Figure 3)

†† Consanguineous family

**Table S2. Clinical features of individuals with *UBA5*-associated DEE44**

(See separate Excel table)

**Table S3. Bioinformatic predictions of the pathogenicity of reported *UBA5* variants**

| | Variant 1 | Variant 2 |
|---|---|---|
| Genomic position (GRCh38) | 3:132665830A>G | 3:132394214A>T |
| Amino acid change | p.Met57Val | p.Gln312Leu |
| Allele frequency in gnomAD | Absent | Absent |
| CADD score | 25.3 | 29.1 |
| SIFT | Damaging | Damaging |
| PolyPhen2 | 1.000 (probably damaging) | 0.999 (probably damaging) |
| MutationTaster | Disease causing | Disease causing |
| PROVEAN | -3.18 (deleterious) | -6.59 (deleterious) |
